# A deterministic epidemic model for the emergence of COVID-19 in China

**DOI:** 10.1101/2020.03.08.20032854

**Authors:** Meng Wang, Jingtao Qi

## Abstract

Coronavirus disease (COVID-19) broke out in Wuhan, Hubei province, China, in December 2019 and soon after Chinese health authorities took unprecedented prevention and control measures to curb the spreading of the novel coronavirus-related pneumonia. We develop a mathematical model based on daily updates of reported cases to study the evolution of the epidemic. With the model, on 95% confidence level, we estimate the basic reproduction number, *R*_0_ = 2.82 ± 0.11, time between March 19 and March 21 when the effective reproduction number becoming less than one, the epidemic ending after April 2 and the total number of confirmed cases approaching 14408 ± 429 on the Chinese mainland excluding Hubei province.

---

A novel coronavirus disease, named COVID-19, broke out in Wuhan, Hubei province, China, in December 2019. On 20 January 2020, the National Health Commission of the People’s Republic of China announced the top-level prevention and control measures against pneumonia, and China determined to curb the spreading of the disease effectively [1, 2]. The current epidemic situation on the Chinese mainland is shown in Figure 1, with data in use listed in Table 1. Checking the curve of cumulative confirmed cases, we can see there is an abrupt change of increasing trend around January 28, being exponential before the time and softened afterwards. A similarity is also observed on the curve of cumulative removed cases (recoveries plus deaths), with delay of a few days. The change indicates the control measures have taken in effect. How to evaluate the effect quantitatively, and in particular, to predict the evolution of the disease would be highly desired.

**Table 1:** COVID-19 situation on the mainland, China.

| Date | Mainland cumulative |  |  | Hubei Province cumulative |  |  | Excluding Hubei |  |  |
| --- | --- | --- | --- | --- | --- | --- | --- | --- | --- |
|  | Confirmed | Recoveries | Deaths | Confirmed | Recoveries | Deaths | I | R | A=I+R |
| 1/20 | 291 | 25 | 6 | 270 | 25 | 6 | 21 | 0 | 21 |
| 1/21 | 440 | 28 | 9 | 375 | 28 | 9 | 65 | 0 | 65 |
| 1/22 | 571 | 28 | 17 | 444 | 28 | 17 | 127 | 0 | 127 |
| 1/23 | 830 | 34 | 25 | 549 | 31 | 24 | 277 | 4 | 281 |
| 1/24 | 1287 | 38 | 41 | 729 | 32 | 39 | 550 | 8 | 558 |
| 1/25 | 1975 | 49 | 56 | 1052 | 42 | 52 | 912 | 11 | 923 |
| 1/26 | 2744 | 51 | 80 | 1423 | 44 | 76 | 1310 | 11 | 1321 |
| 1/27 | 4515 | 60 | 106 | 2714 | 47 | 100 | 1782 | 19 | 1801 |
| 1/28 | 5974 | 103 | 132 | 3554 | 80 | 125 | 2390 | 30 | 2420 |
| 1/29 | 7711 | 124 | 170 | 4586 | 90 | 162 | 3083 | 42 | 3125 |
| 1/30 | 9692 | 171 | 213 | 5806 | 116 | 204 | 3822 | 64 | 3886 |
| 1/31 | 11791 | 243 | 259 | 7153 | 166 | 249 | 4551 | 87 | 4638 |
| 2/1 | 14380 | 328 | 304 | 9074 | 215 | 294 | 5183 | 123 | 5306 |
| 2/2 | 17205 | 475 | 361 | 11177 | 295 | 350 | 5837 | 191 | 6028 |
| 2/3 | 20438 | 632 | 425 | 13522 | 396 | 414 | 6669 | 247 | 6916 |
| 2/4 | 24324 | 892 | 490 | 16678 | 520 | 479 | 7263 | 383 | 7646 |
| 2/5 | 28018 | 1153 | 563 | 19665 | 633 | 549 | 7819 | 534 | 8353 |
| 2/6 | 31161 | 1540 | 636 | 22112 | 817 | 618 | 8308 | 741 | 9049 |
| 2/7 | 34546 | 2050 | 722 | 24953 | 1115 | 699 | 8635 | 958 | 9593 |
| 2/8 | 37198 | 2649 | 811 | 27013 | 1439 | 780 | 8944 | 1241 | 10185 |
| 2/9 | 40171 | 3281 | 908 | 29631 | 1795 | 871 | 9017 | 1523 | 10540 |
| 2/10 | 42638 | 3996 | 1016 | 31728 | 2222 | 974 | 9094 | 1816 | 10910 |
| 2/11 | 44653 | 4740 | 1113 | 33366 | 2639 | 1068 | 9141 | 2146 | 11287 |
| 2/12 | 59804 | 5911 | 1367 | 48206* | 3441 | 1310 | 9071 | 2527 | 11598 |
| 2/13 | 63851 | 6723 | 1380 | 51986 | 3862 | 1318 | 8942 | 2923 | 11865 |
| 2/14 | 66492 | 8096 | 1523 | 54406 | 4774 | 1457 | 8698 | 3388 | 12086 |
| 2/15 | 68500 | 9419 | 1665 | 56249 | 5623 | 1596 | 8386 | 3865 | 12251 |
| 2/16 | 70548 | 10844 | 1770 | 58182 | 6639 | 1696 | 8087 | 4279 | 12366 |
| 2/17 | 72436 | 12552 | 1868 | 59989 | 7862 | 1789 | 7678 | 4769 | 12447 |
| 2/18 | 74185 | 14376 | 2004 | 61682 | 9128 | 1921 | 7172 | 5331 | 12503 |
| 2/19 | 75002<sup>†</sup> | 16157<sup>†</sup> | 2118 | 62457<sup>†</sup> | 10339<sup>†</sup> | 2029 | 6638 | 5907 | 12545 |
| 2/20 | 75891<sup>†</sup> | 18266<sup>†</sup> | 2236 | 63088<sup>†</sup> | 11790<sup>†</sup> | 2144 | 6235 | 6568 | 12803 |
| 2/21 | 76288 | 20659 | 2345 | 63454 | 13557 | 2250 | 5637 | 7197 | 12834 |
| 2/22 | 76936 | 22888 | 2442 | 64084 | 15299 | 2346 | 5167 | 7685 | 12852 |
| 2/23 | 77150 | 24734 | 2592 | 64287 | 16738 | 2495 | 4770 | 8093 | 12863 |
| 2/24 | 77658 | 27323 | 2663 | 64786 | 18854 | 2563 | 4303 | 8569 | 12872 |
| 2/25 | 78064 | 29745 | 2715 | 65187 | 20912 | 2615 | 3944 | 8933 | 12877 |
| 2/26 | 78497 | 32495 | 2744 | 65596 | 23200 | 2641 | 3503 | 9398 | 12901 |
| 2/27 | 78824 | 36117 | 2788 | 65914 | 26403 | 2682 | 3090 | 9820 | 12910 |
| 2/28 | 79251 | 39002 | 2835 | 66337 | 28895 | 2727 | 2699 | 10215 | 12914 |
| 2/29 | 79824 | 41625 | 2870 | 66907 | 31187 | 2761 | 2370 | 10547 | 12917 |
| 3/1 | 80026 | 44462 | 2912 | 67103 | 33757 | 2803 | 2109 | 10814 | 12923 |
| 3/2 | 80151 | 47204 | 2943 | 67217 | 36167 | 2834 | 1788 | 11146 | 12934 |
| 3/3 | 80270 | 49856 | 2981 | 67332 | 38556 | 2871 | 1528 | 11410 | 12938 |
| 3/4 | 80409 | 52045 | 3012 | 67466 | 40479 | 2902 | 1267 | 11676 | 12943 |
| 3/5 | 80552 | 53726 | 3042 | 67592 | 41966 | 2931 | 1089 | 11871 | 12960 |
| 3/6 | 80651 | 55404 | 3070 | 67666 | 43468 | 2959 | 938 | 12047 | 12985 |
| 3/7 | 80695 | 57065 | 3097 | 67707 | 45011 | 2986 | 823 | 12165 | 12988 |

Data are collected from the official web pages of the National Health Commission of the People's Republic of China [3] and cross-checked with reports on the official web pages of the Health Commission of Hubei Province [4]. Data are collected from the official web page of National Health Commission of the People's Republic of China. Only cases on the Chinese mainland have been used in analysis. Numbers in the three rightmost columns are defined in the text. \*Hubei Province added clinically diagnosed cases (only applicable to Hubei province) to laboratory confirmed cases. <sup>†</sup>These numbers are corrected according to reports of Health Commission of Hubei Province.

**Figure 1:**
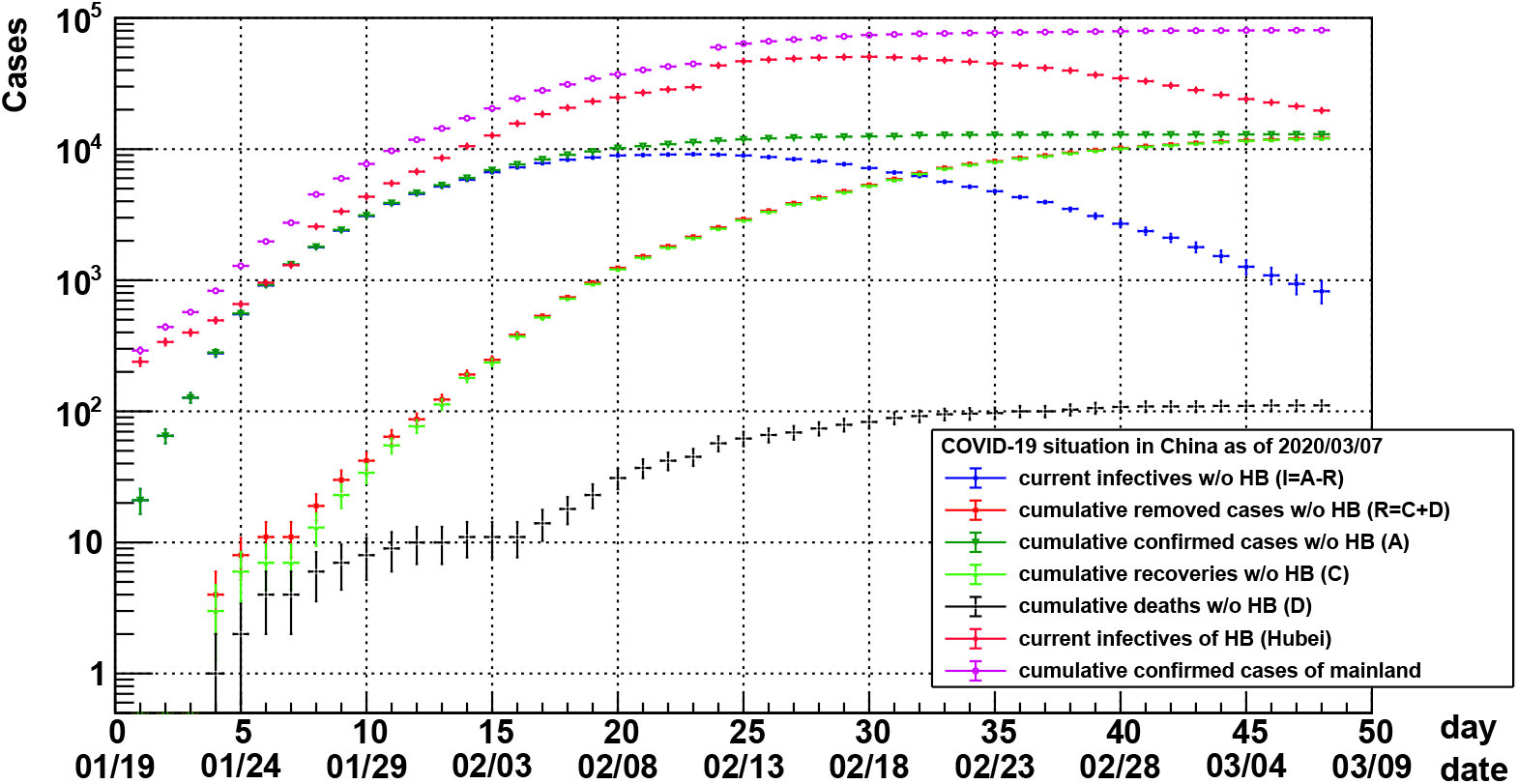
Epidemic curves of reported COVID-19 cases on the Chinese mainland since 20 January 2020. Data are collected from the official web pages of the National Health Commission of the People’s Republic of China [3] and cross-checked with reports on the official web pages of the Health Commission of Hubei Province [4]. In the legend, HB stands for Hubei province, and *I* and *R* represent two categories to be fitted with an mathematical model.

Many epidemic models exist to describe spreads of infectious diseases mathematically [5]. The central role of modelling is to estimate the basic reproduction number, *R*_0_, defined as the expected average number of secondary cases infected by a typical primary case in a fully susceptible population, as well as the effective reproduction number, *R*_*t*_, defined as the actual average number of secondary cases per primary case during an epidemic. Different approaches [6, 7] have been taken for the purpose. A study on COVID-19 in Wuhan [8] is also reported. In rapid development of an infectious disease like COVID-19 in China and in the world, however, these approaches are very difficult in use, if not impossible, or lack of accuracy.

We consider a mathematical model purely based on categorised reported cases, because we believe that key epidemic features will be convolved in the evolution of an infectious disease, namely, the statistics of epidemic data. Should such a model can be fitted with data reasonably well, some epidemic parameters can be extracted. Let’s take the number of cumulative confirmed cases (*A*), the number of cumulative recovered cases (*C*) and the number of cumulative deaths (*D*). We define the number of cumulative removed cases (*R*) as *R* = *C* + *D*, and the number of current infectives (*I*) as *I* = *A− R*. We assume change rates of both *I* and *R* are proportional to *I* (justification of this assumption will been discussed later),

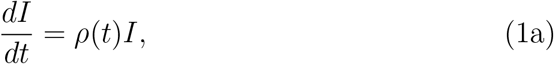

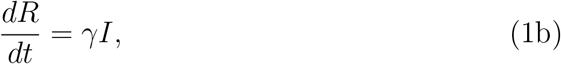

where *ρ* denotes the change rate of *I* and in general varies with time, *γ* is the change rate of *R* and can be regarded as a constant. For the time varying *ρ*, the equation (1a) can be solved analytically

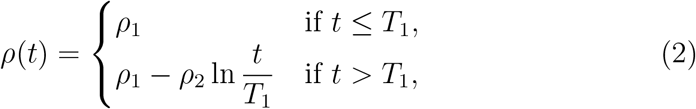

where *T*_1_ is a transition time when the rate changes from a constant to a time dependent variable due to disease control measures taking effect, and both *ρ*_1_ and *ρ*_2_ are constants. Then, we have

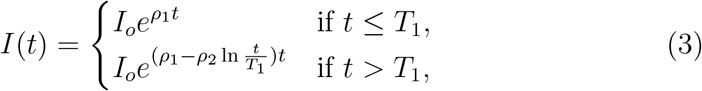

where *I*_*o*_ = *I*(*t* = 0) is a constant. Considering the existence of an infectious period, the transition time of *R*(*t*) will be different from that of *I*(*t*). Therefore, *R*(*t*) can be expressed as

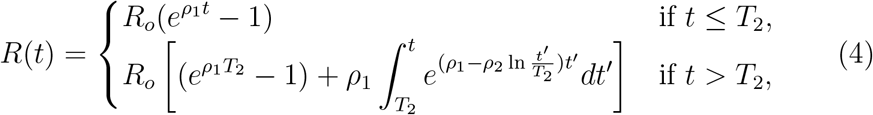

where *T*_2_ is a transition time of *R*, and *R*_*o*_ = *R*(*t* = 0), being a constant. *R*_*o*_ is related to *I*_*o*_ by *R*_*o*_ = *γI*_*o*_*/ρ*_1_. But in principle, both *I*_*o*_ and *R*_*o*_ are just arbitrary constants because we choose time zero randomly.

We use (3) and (4) to fit epidemic curves. For various reasons, reported numbers of cumulative confirmed cases in Hubei are comparatively less reliable. Particularly, there is a jump on February 12, seeing Figure 1 and Table 1, due to the addition of clinically diagnosed cases (only applicable to Hubei province) to previously laboratory confirmed cases. On the other hand, numbers of cumulative confirmed cases, recoveries and deaths of other provinces on the Chinese mainland are quite reliable since January 20 when the National Health Commission started to daily report the epidemic situation. Therefore, we apply the model to the epidemic data excluding Hubei.

We fit, in Figure 1, the curve of current infectives (*I*) with (3) and the curve of cumulative removed cases (*R*) with (4) simultaneously. During the fitting procedure, the parameters *ρ*_1_ and *ρ*_2_ are shared, and others are free to vary. The fit method is minimisation of a chi-square (*χ*^2^) using data errors. The data errors are estimated on the base of single counting uncertainty. Let *A*_*i*_, *C*_*i*_ and *D*_*i*_ separately stand for cumulative confirmed cases, cumulative recoveries and cumulative deaths reported as of *i*-th day. By definition, *I*_*i*_ = *A*_*i*_ *− C*_*i*_ *− D*_*i*_ and *R*_*i*_ = *C*_*i*_ + *D*_*i*_, we have 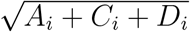 for the error of *I*_*i*_ and 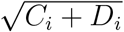 for the error of *R*_*i*_, respectively. Fitting results are shown in Figure 2 and summarised in Table 2. The covariance matrix and the correlation matrix of fit parameters are also reported in Table 3 and Table 4, respectively. The 95% confidence intervals have been calculated for *I, R* as well as *A*, also shown in Figure 2.

**Table 2:** Fitting results.

| Parameter | unit | Value |  |
| --- | --- | --- | --- |
| $\chi^2$ | | 169.992 | |
| ndf |  | 84 |  |
| $I_o$ | | $42.532 \pm 3.22136$ | |
| $T_1$ | day | $9.56175 \pm 0.293452$ | |
| $R_o$ | | $0.51429 \pm 0.0641836$ | |
| $T_2$ | day | $13.6699 \pm 0.421613$ | |
| $\rho_1$ | $\text{day}^{-1}$ | $0.442754 \pm 0.0122409$ | |
| $\rho_2$ | $\text{day}^{-1}$ | $0.237815 \pm 0.00309522$ | |
| $\beta$ | $\text{day}^{-1}$ | $0.686173 \pm 0.0773438$ | (95% CL) |
| $1/\gamma$ | day | $4.10814 \pm 0.544044$ | (95% CL) |
| $R_0$ | | $2.8189 \pm 0.107153$ | (95% CL) |

**Table 3:** Covariance matrix of fit parameters.

| | $I_o$ | $T_1$ | $R_o$ | $T_2$ | $\rho_1$ | $\rho_2$ |
| --- | --- | --- | --- | --- | --- | --- |
| $I_o$ | 10.378 | 0.650837 | 0.194238 | 1.01997 | -0.035529 | -0.00971026 |
| $T_1$ | 0.650837 | 0.0861171 | 0.0153761 | 0.121913 | -0.00334855 | -0.000603745 |
| $R_o$ | 0.194238 | 0.0153761 | 0.00411954 | 0.02227 | -0.000749944 | -0.000187415 |
| $T_2$ | 1.01997 | 0.121913 | 0.02227 | 0.177767 | -0.00491065 | -0.000931429 |
| $\rho_1$ | -0.035529 | -0.00334855 | -0.000749944 | -0.00491065 | 0.000149839 | 3.35947e-05 |
| $\rho_2$ | -0.00971026 | -0.000603745 | -0.000187415 | -0.000931429 | 3.35947e-05 | 9.58036e-06 |

**Table 4:** Correlation matrix of fit parameters.

| | $I_o$ | $T_1$ | $R_o$ | $T_2$ | $\rho_1$ | $\rho_2$ |
| --- | --- | --- | --- | --- | --- | --- |
| $I_o$ | 1 | 0.688446 | 0.939403 | 0.750942 | -0.900978 | -0.973829 |
| $T_1$ | 0.688446 | 1 | 0.816353 | 0.985328 | -0.932181 | -0.664688 |
| $R_o$ | 0.939403 | 0.816353 | 1 | 0.822945 | -0.954537 | -0.943384 |
| $T_2$ | 0.750942 | 0.985328 | 0.822945 | 1 | -0.951486 | -0.713729 |
| $\rho_1$ | -0.900978 | -0.932181 | -0.954537 | -0.951486 | 1 | 0.886681 |
| $\rho_2$ | -0.973829 | -0.664688 | -0.943384 | -0.713729 | 0.886681 | 1 |

**Figure 2:**
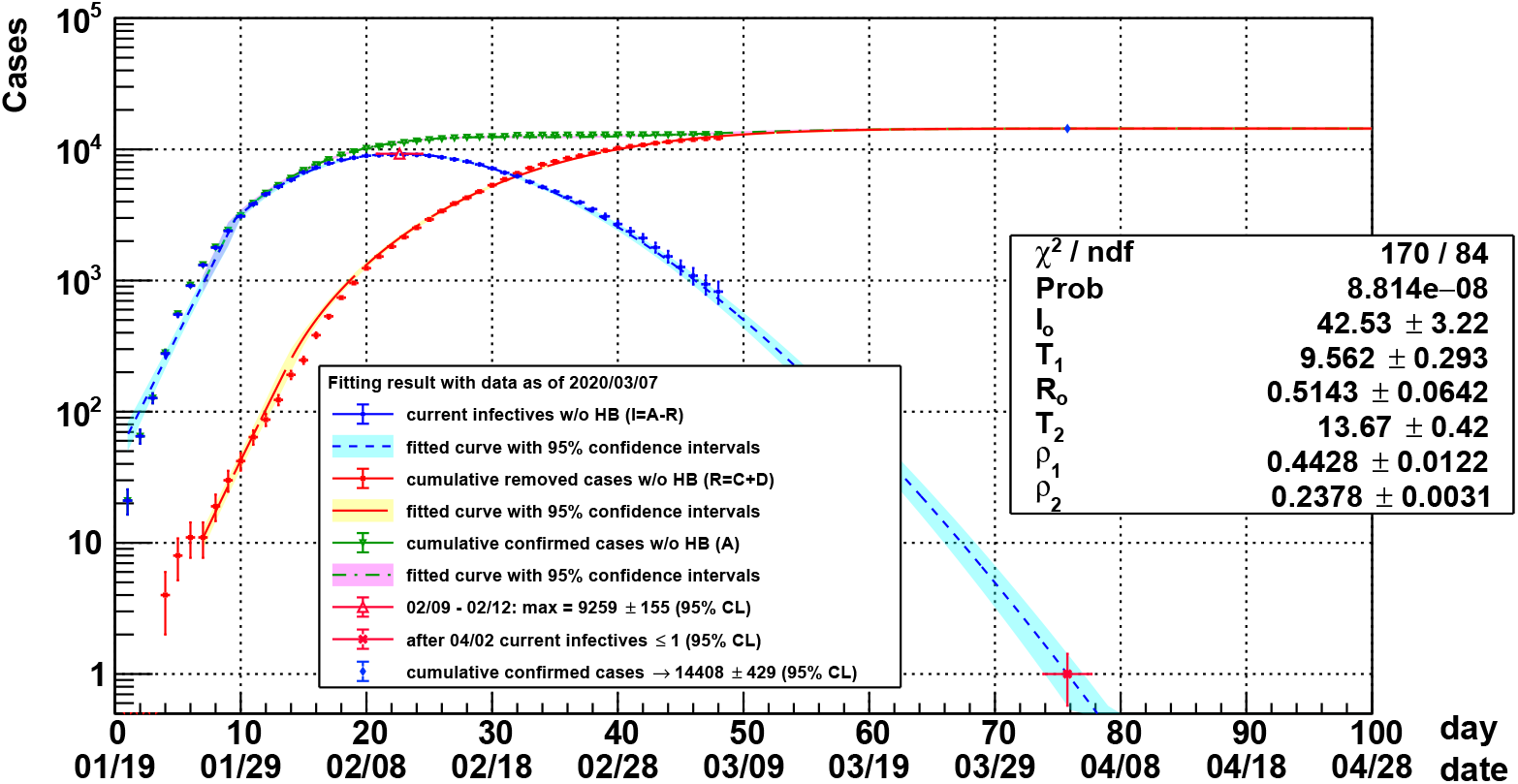
Fitting results of COVID-19 on the Chinese mainland. In the figure, dots with error bars are epidemic data, curves are fitted functions and colourful bands are 95% confidence intervals.

The spreading trend of COVID-19 on the Chinese mainland (excluding Hubei) could be predicted with the model. With the fitting results, the number of current infectives would be less than one after April 2 with 95% confidence level (CL)^1^, that can be regarded as the ending of the disease. Meanwhile, the total number of confirmed cases would approach 14408±429. In addition, we see the current infectives peaks around February 11. Should we perform the same analysis on data before the peaking date, we could obtain similar results, but with comparatively larger uncertainties, as shown in Figure 3, confirming the predictability of the method.

**Figure 3:**
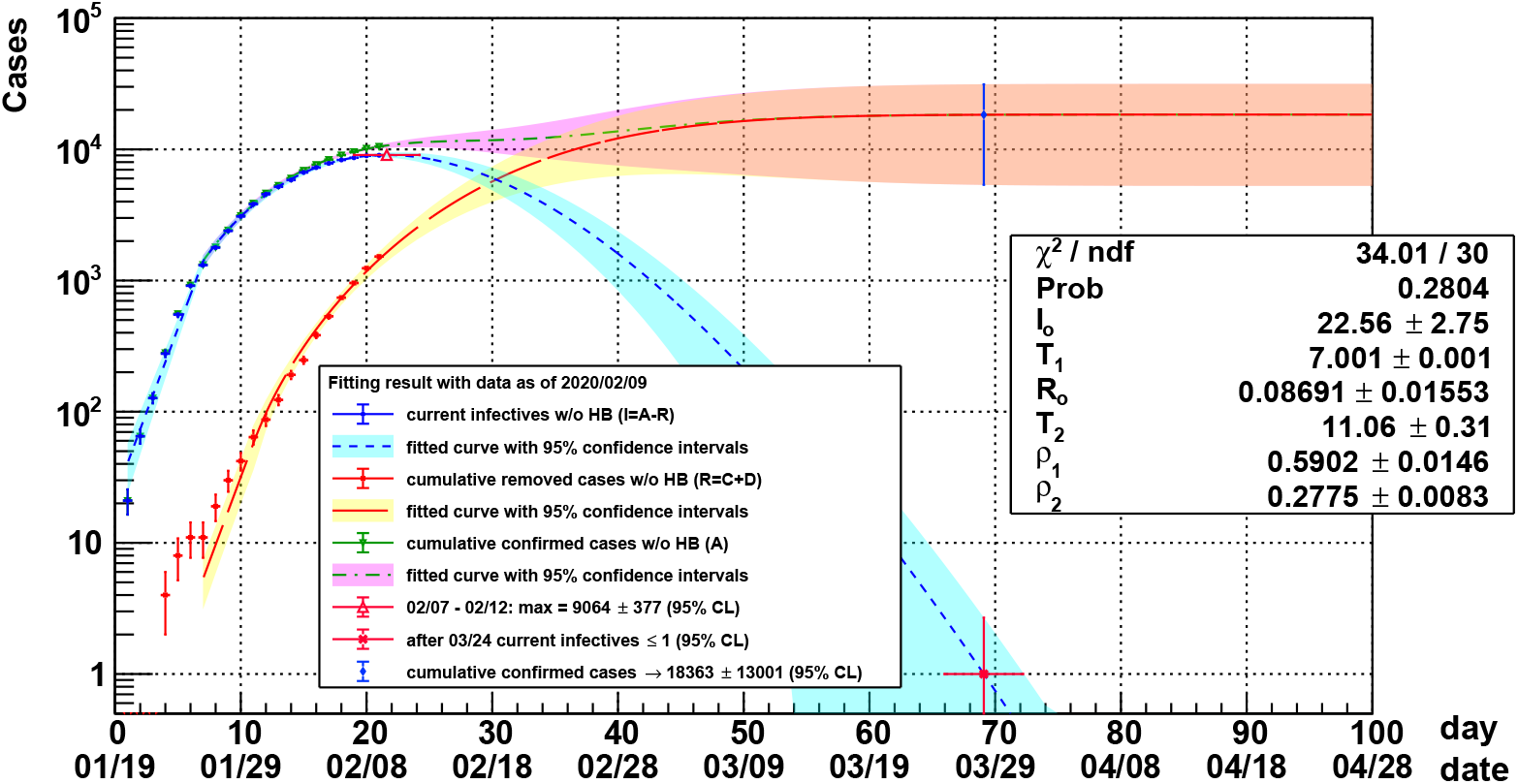
Fitting results of COVID-19 on the Chinese mainland with data as of February 9. In the figure, dots with error bars are epidemic data, curves are fitted functions and colourful bands are 95% confidence intervals.

Our model and results can be compared to the classic Susceptible-InfectiveRemoved (SIR) model. The SIR model divides the population into compartments: the compartment *S* for susceptibles, the compartment *I* for infectives and the compartment *R* for removed (recovered or dead) cases. Corresponding numbers in the compartments are related by a set of derivative equations

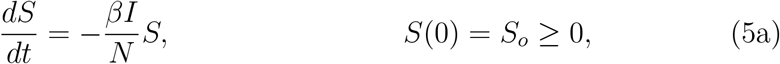

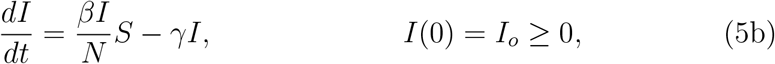

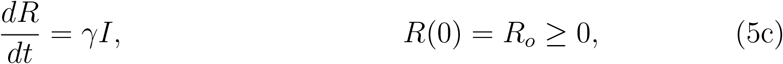

where *N* = *S* + *I* + *R* is the total population, *β* is the average number of contacts sufficient for transmission per person per unit time (contact rate) and 1*/γ* is the average infectious period, and all the three parameters are regarded as constants. The model is adequate for limited population and freely developed infectious diseases. For COVID-19 in China, the static SIR model is incapable. On one hand, the extremely tight prevention and control measures have taken effect significantly, hence suppressing the contact rate with time; on the other hand, the population in China is more than 1.4 billion and modern transportation methods, e.g., high speed trains, have brought all people in the country as a whole being susceptible, eliminating the depletion effect of susceptibles. The latter causes *S ≈ N* can be regarded as being infinite, or a constant. Consequently the equations in (5) reduce to our model in (1) with

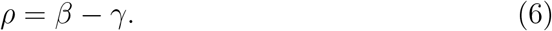

Before the transition time *T*_1_, *ρ*_1_ = *β − γ*, being a constant, justifying our previous assumption for the model. To estimate *γ*, we cannot use *I*_*o*_ and *R*_*o*_ directly because the time zero, *t* = 0, in our analysis is arbitrarily chosen and the two constants are subject to large fluctuation. Instead, we can take the epidemic meaning of 1*/γ* and interpret its equivalence to the transition time difference between *R* and *I*, namely,

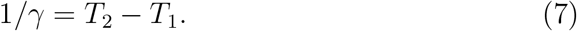

Resolving *β* and *γ*, we can estimate the basic reproduction number

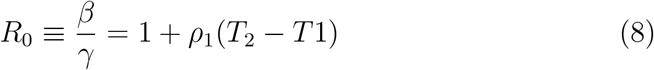

with our fitting results, *R*_0_ = 2.82±0.11 (95% CL).

To quantify the impact of control measures, however, we should use the effective reproduction number, *R*_*t*_. The spreading of an epidemic will stop when *R*_*t*_ become less than 1 persistently. With respect to the time dependent contact rate, *β*(*t*), using (6), we have the effective reproduction number

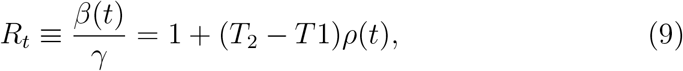

where *ρ*(*t*) is in (2). The distribution of *R*_*t*_ with our fitting results is shown in Figure 4. It starts with *R*_0_, and approaches zero logarithmically. The time when *R*_*t*_ = 1 will be between March 19 and March 21 (95% CL).

**Figure 4:**
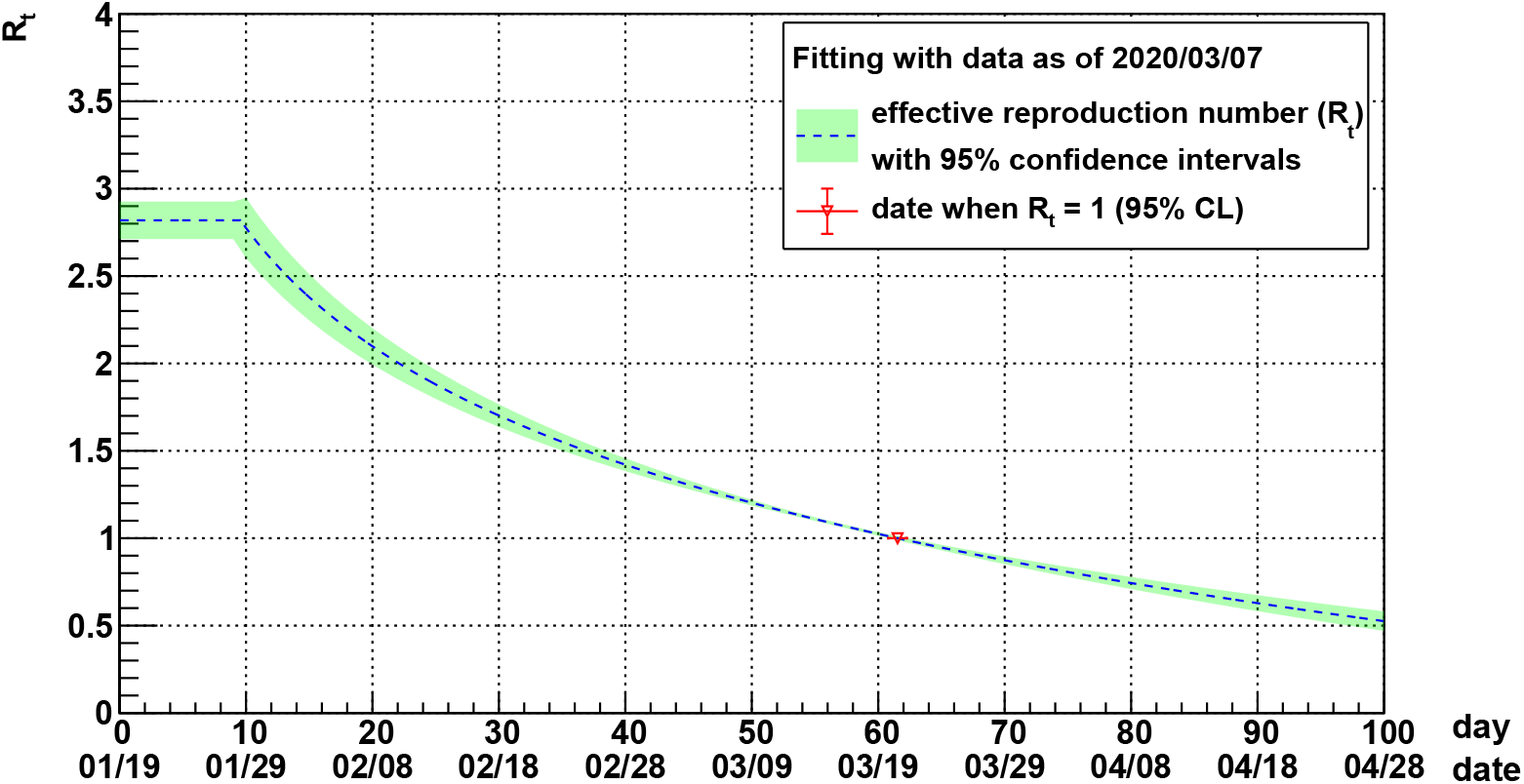
The effective reproduction number, *R*_*t*_, deduced from fitting results. The predicted date when *R*_*t*_ = 1 is indicated on the curve.

The stability and consistency of our model can be verified by fitting the previous epidemic data day-by-day up to now. The repetitious fit starts with data as of February 5 in order to have enough statistics. For each fit, we extract *R*_0_ and the date, 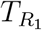, with *R*_*t*_ = 1. Results are depicted in Figure 5. We see fluctuation in daily results of *R*_0_ and 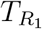, but both of them are consistent within 95% confidence intervals during the epidemic so far.

**Figure 5:**
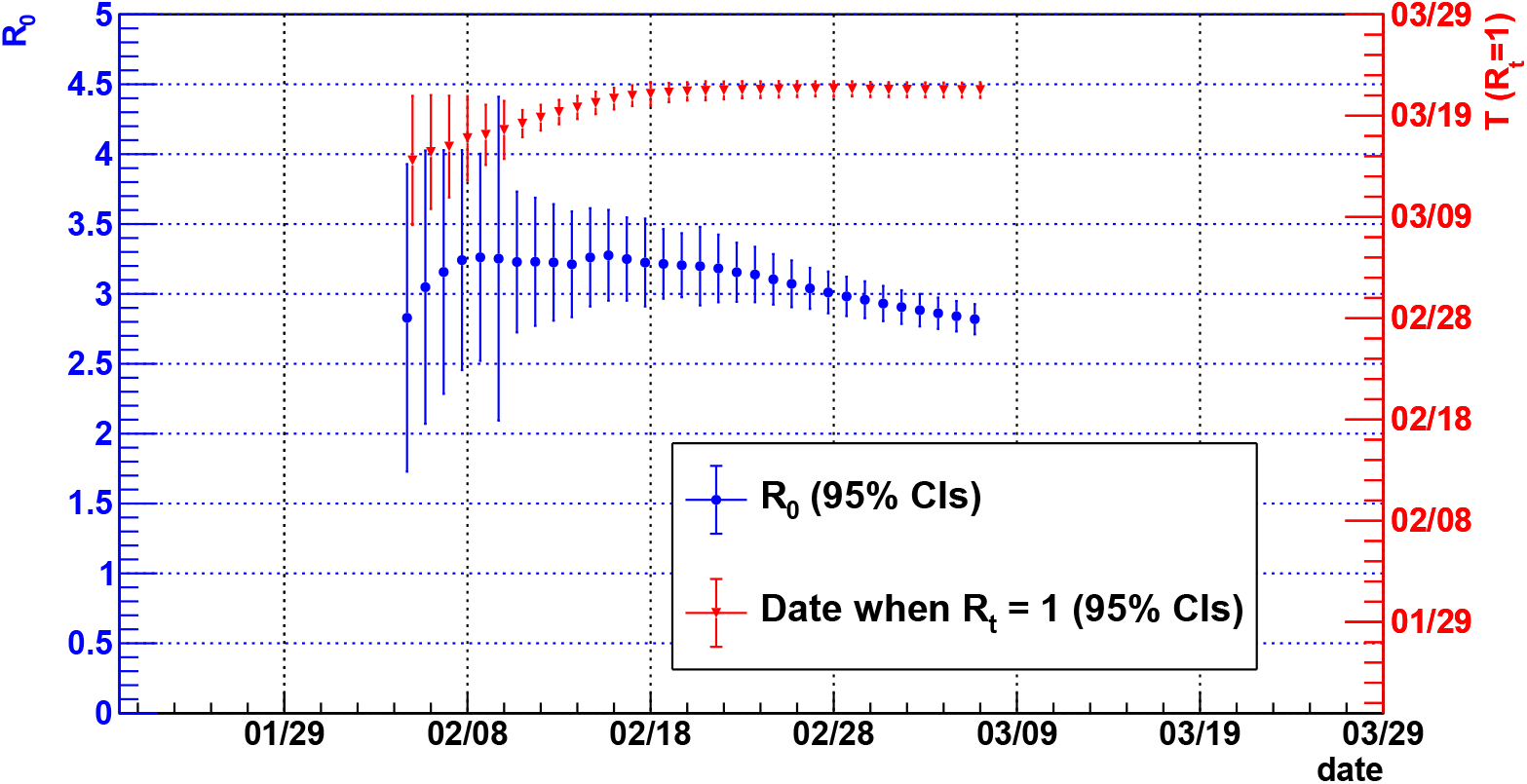
Estimates of the basic reproduction number (*R*_0_) and the predicted date 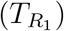 when the effective reproduction number (*R*_*t*_) reaching one as functions of date of situation report.

## Discussion

The study provides a deterministic mathematical model to analyse and predict characteristics of an infectious disease spreading in an extremely large population with strong prevention and control measures. The model depends purely on statistic data of categorised epidemic cases reported in public. We apply the model to the emergence of COVID-19 in China. Although the model is not yet perfect, it is effective, robust and consistent as long as there is no sudden change in control measures. Comparing with the classic SIR model, we extract the basic reproduction number, *R*_0_, and make predictions on the time when the effective reproduction number will become less than one and on the total number of confirmed cases in the end. Our study indicates that it is essential to take effective control measures as early as possible for curbing an infectious disease and it is important to report epidemic data precisely and timely for epidemiological modelling. In the era of globalisation, an infectious disease can spread as far as any individual can reach on the earth, that makes the whole world population to be susceptible. As COVID-19 has spread around the world, we anticipate our method can be adapted in modelling the spread and help public health authorities make policy decisions.

## Data Availability

The epidemic data are collected from reports on the official web pages of National Health Commission of the People's Republic of China.

http://www.nhc.gov.cn/xcs/yqtb/list_gzbd.shtml

## Footnotes

1 All uncertainties of predictions in the paper are reported on 95% confidence level.

## Notes

### Competing Interest Statement

The authors have declared no competing interest.

### Funding Statement

none.

